## Supplementary Material for "Social mixing patterns in the UK following the relaxation of COVID-19 pandemic restrictions: a cross-sectional online survey"

### Analysis

#### Age-specific mixing rates

To calculate age-specific mixing patterns we first defined matrix $C_{ij}$, where $C_{ij}$ was the number of non-household contacts reported between participant age groups *i* and contact age groups *j*. The mean contact rate per age group ($M_{ij}$) was given by

$$M_{ij} = \frac{C_{ij}}{N_i}$$

where $N_i$ was the number of participants in age group *i*. As a measure of uncertainty, we calculated confidence intervals by taking 1,000 bootstrapped samples of participants.

Similarly, age-specific non-household contact rates were derived from the POLYMOD data.[1] We calculated the percentage decrease of age-specific non-household contact rates between the POLYMOD data and the CoCoNet data; Figure S1.

We found moderate assortative mixing by age, in line with both current and pre-pandemic contact studies (q = 0.38, 95%CI 0.17 to 0.58); Figure S1. Of all ages under 80 years old, 30-39 year olds had the highest non-household contact rate with those aged 80 or over; 4.8% (95%CIs 3.77 to 6.04) of non-household contacts reported by 30-39 years olds were with someone aged 80+.

#### Variables associated with variation in non-household contact rate - model selection

To identify characteristics of the participant associated with their rate of daily non-household contact we fitted a negative binomial model to the daily number of non-household contacts reported by participants. We then used our selected explanatory variables as candidate variables for a forward stepwise model selection process.

Participant age group, sex, ethnicity, working situation and COVID circumstance were included as explanatory variables in all models. Candidate explanatory variables were: nation of residence (England, Northern Ireland, Scotland or Wales); household size; dwelling type; whether the contact reporting day was a weekend or week day; whether the participant had left their home on the contact reporting day.

The model selection process selected the following variables from candidate variables: age, sex, ethnicity, working situation, COVID circumstance, nation, household size, day of the week, whether the participant had left their home as explanatory variables. Dwelling type was not selected as an explanatory variable. Their association with the rate of non-household contact in the fully adjusted model is shown in Table S7.

#### Contact clustering or transitivity

Participants who made fewer than 15 contacts and did not live alone were asked if anyone in their household had met each of their contacts that same day, as a measure of clustering (also called transitivity) within social networks.[2] We estimated 40.4% (95%CI 39.1 to 41.6) of non-household contacts were also met by another household member on the same day. The proportion of contacts encountered by participant and household members was highest if the contact was under 20 years old: contact aged 0 to 4 (77.5%, 95%CI 66.8 to 86.1); aged 5 to 9 (80.0%, 95%CI 72.7 to 86.1); aged 10 to 19 (63.2%, 95%CI 56.1 to 69.9). This indicates that non-household interactions with children tend to be made with multiple individuals from those households.

**Visiting other households**

Evidence suggests that transmission of COVID-19 often occurs within households.[3] We found that 12.2% (95%CI 11.3 to 13.1) of participants reported visiting another household. Females (aOR 1.2, 95%CI 0.98 to 1.56) and members of support bubbles (aOR 1.92, 95%CI 1.61 to 2.28) were more likely to have visited another household; Table S8.

**Household visits**

The mean rate of contacts made with non-household members (non-household contacts) by those not leaving their home was 0.4 $d^{-1}$ (95%CI 0.4 to 0.5), with 23.8% (95%CI 21.2 to 26.6) meeting one or more non-household contacts. This contact rate was significantly lower than for those who did leave their home (incidence rate ratio (IRR) 0.12, 95%CI 0.11 to 0.14, p-value <0.001).

**Support bubbles**

A substantial proportion (40.1%, 95%CI 38.7 to 41.4) of participants reported being part of a support bubble, with 43.1% (95%CI 40.9 to 45.3) joining with a single-person household. Males were less likely to be part of a support bubble (aOR 0.68, 95%CI 0.58 to 0.78); support bubble membership was not associated with age group or ethnicity. Support bubbles had a median (non-participant side) size of 2.0 (25th percentile 1.0, 75th percentile 3.0) and mean size of 2.2, (95%CI 2.1 to 2.2), and were mostly encountered two or fewer days in the past week.

**Survey methodology - limitations**

When calculating the mean number of daily non-household contacts, an assumption for the maximum number of contacts was made. The survey asked participants how many contacts they had made yesterday, with the option of '0 to 15 or more'. If participants selected '15 or more', they were asked to group the contacts they had made by age, by selecting an integer between 0 and 19 or '20+' for each contact age category. To calculate the mean number of contacts, where '20+' contacts was selected, this was assumed to be 20 contacts. We may, therefore, have underestimated the maximum number of contacts of some participants and non-household contact rates.

### Figures

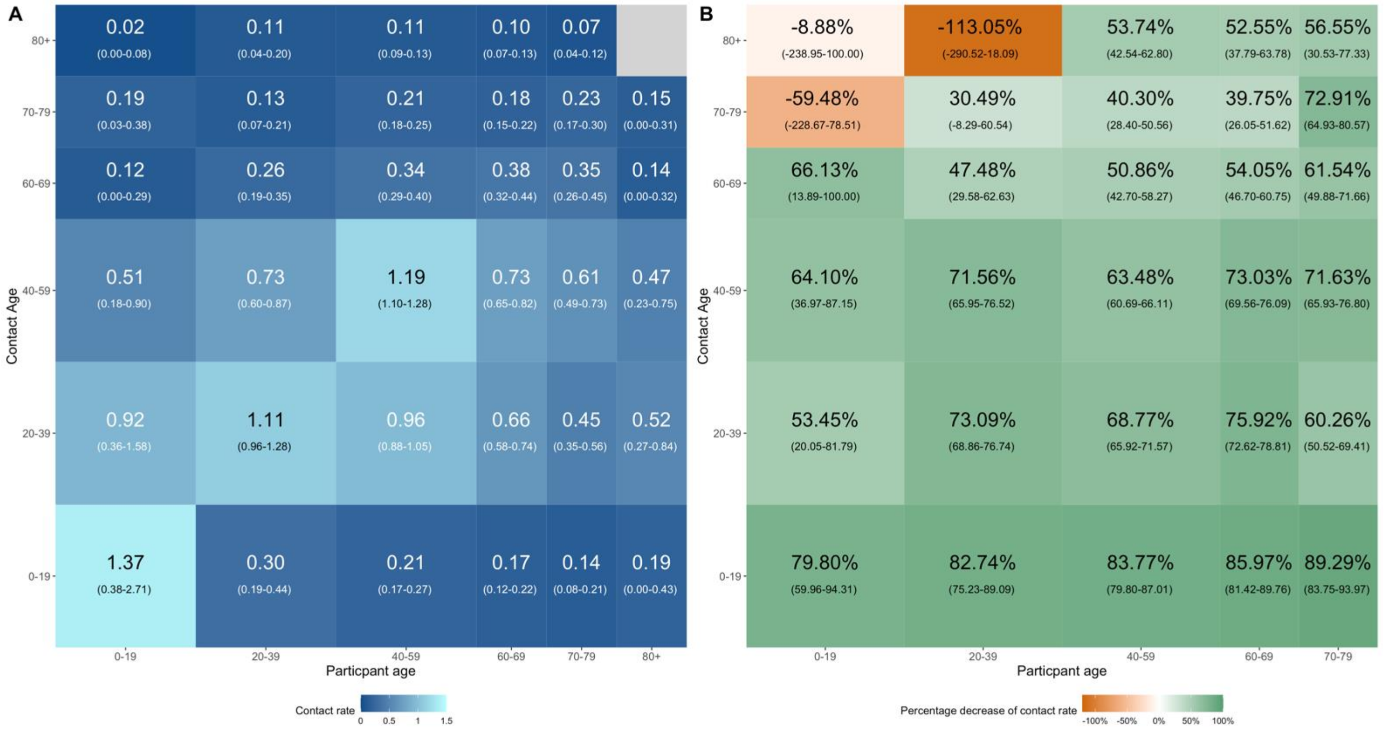

**Figure S1.** (A) Mean non-household contact rate (number of contacts per day) with different age groups reported by participant age group; bootstrapped 95% confidence intervals shown in parentheses. (B) Percentage decrease of non-household contact rate between the POLYMOD data (2005-2006) and the CoCoNet data; bootstrapped 95% confidence intervals shown in parentheses.

**Tables**

**Table S2.** Comparison of non-household contact rates across different UK contact surveys.

| | Mean contact rate, $d^{-1}$ (25th and 75th percentile) | | | | |
|---|---|---|---|---|---|
| Study, contact type and geography of sample | **CoCoNet Number of participants** | **CoCoNet non-household contacts UK** | **CoMix non-household contacts UK** [4] | **Social Contact Survey non-household contacts, Great Britain** [5] | **POLYMOD, non-household contacts, Great Britain** [1] |
| Sampling period | | 28 July 2020 to 14 August 2020 Non-lockdown period | 24 March 2020 to 27 March 2020 Lockdown period | 2009 Pre-pandemic | 2005-2006 Pre-pandemic |
| All participants | 5,037 | 2.9 (0, 3) | 1.4 (0, 1) | 25.9 (5, 23) | 9.6 (4, 13) |
| Age group | | | | | |
| 0-9 | 5 | 1.0 (0, 1) | - | 29.8 (7, 46) | 8.9 (3, 13) |
| 10-19 | 37 | 3.6 (0, 3) | - | 43.1 (8, 40) | 12.3 (5, 18) |
| 20-29 | 250 | 3.2 (0, 3) | 1.1 (0, 1)* | 29.0 (7, 27) | 9.7 (5, 13) |
| 30-39 | 594 | 2.6 (0, 3) | 1.4 (0, 1) | 25.4 (6, 25) | 8.9 (4, 12) |
| 40-49 | 1167 | 3.1 (0, 4) | 1.4 (0, 1) | 30.8 (6, 29) | 9.8 (4, 13) |
| 50-59 | 1617 | 3.3 (0, 3) | 1.6 (0, 2) | 28.6 (6, 26) | 8.2 (3, 11) |
| 60-69 | 1056 | 2.3 (0, 3) | 1.4 (0, 2) | 23.0 (4, 18) | 8.2 (4, 11) |
| 70-79 | 290 | 1.9 (0, 3) | 1.1 (1, 1)** | 19.1 (3, 17) | 6.8 (3, 11) |
| 80+ | 21 | 1.7 (0, 2) | - | 13.2 (1,10) | - |
| Sex† | | | | | |
| Female | 3978 | 2.8 (0, 3) | 1.4 (0, 1) | 27.5 (5, 26) | 10.2 (4, 14) |
| Male | 1037 | 2.9 (0, 3) | 1.3 (0, 1) | 22.6 (4, 19) | 9.0 (4, 13) |
| Prefer not to say | 22 | 1.5 (0, 2) | - | - | - |
| Household size | | | | | |
| 1 | 875 | 2.8 (0, 3) | 1.6 (1, 2) | 24.3 (4, 20) | 7.5 (3, 11) |
| 2 | 1902 | 2.6 (0, 3) | 1.5 (0, 2) | 23.7 (5, 21) | 9.2 (4, 12) |
| 3 | 979 | 3.2 (0, 3) | 1.2 (0, 1) | 24.6 (5, 25) | 9.6 (4, 14) |
| 4 | 896 | 2.7 (0, 3) | 1.4 (0, 1) | 33.4 (6, 33) | 10.0 (4, 14) |
| 5 | 284 | 3.2 (0, 4) | 1.1 (0, 1) | 30.7 (7, 30) | 10.9 (5, 15) |
| 6+ | 101 | 3.8 (0, 4) | 1.1 (0, 1) | 45.6 (8, 36) | 10.1 (5, 15) |

* CoMix age group 18-29

** CoMix age group 70+

† Comix and POLYMOD report participants' gender rather than sex

**Table S3.** Adjusted incidence rate ratios for number of daily non-household contacts by select variables. Intercept of 0.41 (95 %CI 0.35-0.48). Dispersion parameter of 1.07 (95%CI 1.00-1.14)

| | **Multivariable analysis[1]** | |
|---|---|---|
| | **aIRR (95%CI)** | **p-value** |
| Age | | |
| 0-9 | 0.70 (0.13-3.69) | 0.673 |
| 10-19 | 0.90 (0.50-1.67) | 0.724 |
| 20-29 | 1.16 (0.97-1.39) | 0.109 |
| 30-39 | 0.86 (0.76-0.97) | 0.016 |
| 40-49 | 0.90 (0.82-1.00) | 0.039 |
| 50-59 | 1.00 | - |
| 60-69 | 0.89 (0.79-1.00) | 0.046 |
| 70-79 | 0.94 (0.78-1.14) | 0.542 |
| 80+ | 1.23 (0.69-2.25) | 0.458 |
| Sex | | |
| Female | 1.00 | - |
| Male | 1.01 (0.93-1.10) | 0.772 |
| Prefer not to say | 0.53 (0.27-1.05) | 0.063 |
| Ethnicity | | |
| White | 1.00) | - |
| Mixed/Multiple ethnic groups | 1.09 (0.77-1.58) | 0.625 |
| Asian/Asian British | 0.54 (0.36-0.82) | 0.004 |
| Black/African/Caribbean/Black British | 0.46 (0.17-1.23) | 0.136 |
| Other ethnic group | 0.54 (0.18-1.80) | 0.308 |
| Nation | | |
| England | 1.00 | - |
| Northern Ireland | 0.92 (0.59-1.47) | 0.713 |
| Scotland | 0.80 (0.68-0.95) | 0.009 |
| Wales | 1.22 (0.99-1.50) | 0.060 |
| Household size | | |
| 1 | 1.00 | - |
| 2 | 0.92 (0.83-1.02) | 0.105 |

| | Multivariable analysis[1] | |
|---|---|---|
| | aIRR (95%CI) | p-value |
| 3 | 0.99 (0.88-1.12) | 0.878 |
| 4 | 0.84 (0.74-0.96) | 0.008 |
| 5 | 0.97 (0.81-1.16) | 0.717 |
| 6+ | 0.85 (0.65-1.12) | 0.247 |
| Day of the week (contacts recorded) | | |
| Weekday | 1.00 | - |
| Weekend | 1.14 (0.99-1.31) | 0.066 |
| Left home yesterday | | |
| No | 1.00 | - |
| Yes | 5.58 (4.92-6.33) | <0.001 |
| Dwelling type | | |
| House or bungalow | 1.00 | - |
| Flat, maisonette or apartment | 0.95 (0.85-1.07) | 0.435 |
| Mobile or temporary structure | 1.13 (0.53-2.65) | 0.760 |
| Assisted living facility | 2.25 (0.39-37.68) | 0.442 |
| Care home | 2.84 (0.42-50.22) | 0.345 |
| Other | 1.28 (0.68-2.54) | 0.461 |
| School or work situation | | |
| Employed - working from home | 1.00 | - |
| School pupil - studying at home | 0.83 (0.28-2.46) | 0.739 |
| School pupil - attending school | 2.90 (0.95-10.02) | 0.080 |
| College or University student | 0.84 (0.63-1.12) | 0.229 |
| Employed - going to place of work | 3.33 (3.02-3.66) | <0.001 |
| Self Employed | 1.63 (1.43-1.87) | <0.001 |
| Healthcare professional | 5.10 (4.29-6.10) | <0.001 |
| Unemployed | 1.10 (0.89-1.36) | 0.399 |
| Furloughed | 1.20 (0.98-1.48) | 0.074 |
| Unable to work | 0.96 (0.71-1.31) | 0.815 |
| Retired | 1.24 (1.09-1.42) | 0.001 |
| COVID-19 circumstance | | |
| Not self isolating or shielding | 1.00 | - |

| | Multivariable analysis[1] | |
|---|---|---|
| | aIRR (95%CI) | p-value |
| Self isolating - I have symptoms | 4.05 (1.94-9.72) | 0.001 |
| Self isolating - Someone in my household has symptoms | 1.31 (0.51-3.63) | 0.604 |
| Self isolating - Someone in my support bubble has symptoms | 0.81 (0.21-3.32) | 0.771 |
| Self isolating - precaution/told to by Test and Trace | 0.58 (0.43-0.79) | <0.001 |
| Shielding - I am a vulnerable individual | 0.82 (0.66-1.01) | 0.056 |
| Shielding - I live with a vulnerable individual | 0.79 (0.62-1.02) | 0.065 |
| Not sure | 0.56 (0.39-0.79) | 0.001 |

[1]Adjusted for age,sex, ethnicity, nation, household size, day of the week, left home, dwelling type, school or work situation and COVID-19 circumstance.
*This increased contact rate is due to one participant who was self-isolating with symptoms reporting a large number of contacts (*see results*).

**Table S4.** Characteristics of participants who reported 'Self isolating' or 'Shielding' as their COVID circumstance. N is the number of participants who provided a response to the question.

| | Number of self-isolating participants (%) | Number of shielding participants (%) | Number of participants not self-isolating or shielding (%) |
|---|---|---|---|
| Age group | N = 136 | N = 353 | N = 4,511 |
| 0-9 | 0 (0.0%) | 0 (0.0%) | 5 (0.1%) |
| 10-19 | 1 (0.7%) | 4 (1.1%) | 33 (0.7%) |
| 20-29 | 7 (5.1%) | 12 (3.4%) | 233 (5.2%) |
| 30-39 | 11 (8.1%) | 28 (7.9%) | 549 (12.2%) |
| 40-49 | 17 (12.5%) | 64 (18.1%) | 1093 24.2%) |
| 50-59 | 41 (30.1%) | 95 (26.9%) | 1465 (32.5%) |
| 60-69 | 32 (23.5%) | 104 (29.5%) | 905 (20.1%) |
| 70-79 | 22 (16.2%) | 43 (12.2%) | 215 (4.8%) |
| 80+ | 5 (3.7%) | 3 (0.8%) | 13 (0.3%) |
| No response | 0 (0.0%) | 0 (0.0%) | 0 (0.0%) |
| Sex | N = 136 | N = 353 | N = 4,511 |
| Female | 111 (81.6%) | 286 (81.0%) | 3548 (78.7%) |
| Male | 25 (18.4%) | 65 (18.4%) | 943 (20.9%) |
| Prefer not to say | 0 (0.0%) | 2 (0.6%) | 20 (0.4%) |
| No response | 0 (0.0%) | 0 (0.0%) | 0 (0.0%) |
| Ethnicity | N = 136 | N = 353 | N = 4,511 |
| White | 126 (92.6%) | 334 (94.6%) | 4336 (96.1%) |
| Mixed/Multiple ethnic groups | 5 (3.7%) | 8 (2.3%) | 36 (0.8%) |
| Asian/Asian British | 3 (2.2%) | 2 (0.6%) | 43 (1.0%) |
| Black/African/Caribbean/Black British | 0 (0.0%) | 2 (0.6%) | 9 (0.2%) |
| Other ethnic group | 1 (0.7%) | 0 (0.0%) | 6 (0.1%) |
| Prefer not to say | 0 (0.0%) | 3 (0.8%) | 13 (0.3%) |
| No response | 1 (0.7%) | 4 (1.1%) | 68 (1.5%) |
| Left home yesterday | N = 135 | N = 350 | N = 4,495 |
| No | 64 (47.4%) | 145 (41.4%) | 778 (17.3%) |
| Yes | 71 (52.6%) | 205 (58.6%) | 3717 (82.7%) |

| | Number of self-isolating participants (%) | Number of shielding participants (%) | Number of participants not self-isolating or shielding (%) |
|---|---|---|---|
| No response | 0 (0.0%) | 0 (0.0%) | 0 (0.0%) |
| Part of a support bubble | N = 136 | N= 352 | N = 4,505 |
| No | 87 (64.0%) | 238 (67.6%) | 2664 (59.1%) |
| Yes | 49 (36.0%) | 114 (32.4%) | 1841 (40.9%) |
| No response | 0 (0.0%) | 0 (0.0%) | 0 (0.0%) |
| Social distancing | N = 53 | N= 167 | N = 2,989 |
| Yes, all of the time | 39 (73.6%) | 117 (70.1%) | 1728 (57.8%) |
| More than half of the time | 10 (18.9%) | 36 (21.6%) | 877 (29.3%) |
| Less than half of the time | 4 (7.5%) | 11 (6.6%) | 278 (9.3%) |
| No, none of the time | 0 (0.0%) | 3 (1.8%) | 86 (2.9%) |
| Not sure | 0 (0.0%) | 0 (0.0%) | 20 (0.7%) |
| No response | 0 (0.0%) | 0 (0.0%) | 0 (0.0%) |
| | **Mean daily non-household contact rate (IQR)** | | |
| | **Sef-isolating participants** | **Shielding participants** | **Participants not shielding or self-isolating** |
| | N = 134 | N = 348 | N = 4,484 |
| All participants | 1.2 (0, 2) | 1.3 (0, 2) | 3.1 (0, 3) |

**Table S5.** Adjusted incidence rate ratios for number of daily non-household contacts by select variables. Self-isolating individual with large number of contacts removed for this analysis (see results). Intercept of 0.41 (95 %CI 0.35-0.48). Dispersion parameter of 1.07 (95%CI 1.00-1.14)

| | **Multivariable analysis[1]** | |
|---|---|---|
| | **aIRR (95%CI)** | **p-value** |
| Age | | |
| 0-9 | 0.69 (0.13-3.64) | 0.662 |
| 10-19 | 0.89 (0.49-1.64) | 0.691 |
| 20-29 | 1.15 (0.97-1.39) | 0.114 |
| 30-39 | 0.86 (0.76-0.98) | 0.021 |
| 40-49 | 0.91 (0.82-1.00) | 0.044 |
| 50-59 | 1.00 | - |
| 60-69 | 0.89 (0.79-1.00) | 0.047 |
| 70-79 | 0.94 (0.78-1.14) | 0.540 |
| 80+ | 1.23 (0.69-2.25) | 0.457 |
| Sex | | |
| Female | 1.00 | - |
| Male | 1.02 (0.93-1.11) | 0.730 |
| Prefer not to say | 0.53 (0.27-1.05) | 0.063 |
| Ethnicity | | |
| White | 1.00) | - |
| Mixed/Multiple ethnic groups | 1.13 (0.79-1.62) | 0.511 |
| Asian/Asian British | 0.54 (0.36-0.82) | 0.004 |
| Black/African/Caribbean/Black British | 0.47 (0.17-1.23) | 0.137 |
| Other ethnic group | 0.54 (0.18-1.79) | 0.305 |
| Nation | | |
| England | 1.00 | - |
| Northern Ireland | 0.92 (0.59-1.47) | 0.713 |
| Scotland | 0.80 (0.68-0.95) | 0.009 |
| Wales | 1.22 (1.00-1.50) | 0.059 |
| Household size | | |
| 1 | 1.00 | - |

| | Multivariable analysis[1] | |
|---|---|---|
| | aIRR (95%CI) | p-value |
| 2 | 0.92 (0.83-1.02) | 0.105 |
| 3 | 0.98 (0.87-1.11) | 0.805 |
| 4 | 0.84 (0.74-0.95) | 0.007 |
| 5 | 0.97 (0.81-1.15) | 0.692 |
| 6+ | 0.85 (0.65-1.12) | 0.244 |
| Day of the week (contacts recorded) | | |
| Weekday | 1.00 | - |
| Weekend | 1.14 (0.99-1.31) | 0.068 |
| Left home yesterday | | |
| No | 1.00 | - |
| Yes | 5.54 (4.89-6.29) | <0.001 |
| Dwelling type | | |
| House or bungalow | 1.00 | - |
| Flat, maisonette or apartment | 0.95 (0.85-1.07) | 0.414 |
| Mobile or temporary structure | 1.21 (0.56-2.87) | 0.647 |
| Assisted living facility | 2.25 (0.39-37.46) | 0.441 |
| Care home | 2.78 (0.41-48.81) | 0.355 |
| Other | 1.28 (0.68-2.54) | 0.460 |
| School or work situation | | |
| Employed - working from home | 1.00 | - |
| School pupil - studying at home | 0.85 (0.29-2.50) | 0.763 |
| School pupil - attending school | 2.90 (0.95-10.00) | 0.080 |
| College or University student | 0.86 (0.65-1.15) | 0.291 |
| Employed - going to place of work | 3.34 (3.04-3.68) | <0.001 |
| Self Employed | 1.64 (1.43-1.87) | <0.001 |
| Healthcare professional | 5.12 (4.31-6.12) | <0.001 |
| Unemployed | 1.10 (0.89-1.37) | 0.369 |
| Furloughed | 1.21 (0.99-1.49) | 0.063 |
| Unable to work | 1.00 (0.74-1.35) | 0.993 |
| Retired | 1.25 (1.09-1.42) | 0.001 |
| COVID-19 circumstance | | |

| | Multivariable analysis[1] | |
|---|---|---|
| | aIRR (95%CI) | p-value |
| Not self isolating or shielding | 1.00 | - |
| Self isolating - I have symptoms | 0.83 (0.24-2.82) | 0.775 |
| Self isolating - Someone in my household has symptoms | 1.31 (0.51-3.62) | 0.606 |
| Self isolating - Someone in my support bubble has symptoms | 0.81 (0.21-3.30) | 0.768 |
| Self isolating - precaution/told to by Test and Trace | 0.58 (0.43-0.79) | <0.001 |
| Shielding - I am a vulnerable individual | 0.81 (0.66-1.00) | 0.050 |
| Shielding - I live with a vulnerable individual | 0.79 (0.62-1.02) | 0.065 |
| Not sure | 0.56 (0.39-0.79) | 0.001 |

[1] Adjusted for age,sex, ethnicity, nation, household size, day of the week, left home, dwelling type, school or work situation and COVID-19 circumstance.

**Table S6.** Association of participant characteristics and maintaining social distancing more than half of the time with contacts (adjusted odds ratios). N = 3058. Model intercept of 9.34 (7.24-12.20).

| | Adjusted Odds Ratio (95%CI)[1] | p-value |
|---|---|---|
| Age | | |
| 10-19 | 0.30 (0.08-1.25) | 0.077 |
| 20-29 | 0.62 (0.38-1.06) | 0.069 |
| 30-39 | 0.66 (0.46-0.95) | 0.024 |
| 40-49 | 0.88 (0.65-1.19) | 0.415 |
| 50-59 | 1.00 | - |
| 60-69 | 0.88 (0.61-1.28) | 0.496 |
| 70-79 | 0.91 (0.49 -1.79) | 0.778 |
| 80+ | 0.28 (0.08-1.30) | 0.064 |
| School or work situation | | |
| Employed - working from home | 1.00 | - |
| College or University student | 0.68 (0.33-1.55) | 0.335 |
| Employed - going to place of work | 0.71 (0.53-0.96) | 0.025 |
| Self Employed | 1.48 (0.92-2.51) | 0.126 |
| Healthcare professional | 0.26 (0.17-0.40) | <0.001 |
| Unemployed | 0.59 (0.35-1.07) | 0.068 |
| Furloughed | 1.01 (0.54-2.05) | 0.979 |
| Unable to work | 1.58 (0.56-6.61) | 0.453 |
| Retired | 1.40 (0.90-2.19) | 0.136 |

[1] Adjusted for age and school or work situation
* School pupils excluded from analysis due to insufficient data.

**Table S7.** Adjusted incidence rate ratios for number of daily non-household contacts by select variables. Model intercept was 0.40 (95 %CI 0.34-0.47). Dispersion parameter of 1.07 (95%CI 1.00-1.13)

| | **Multivariable analysis[1]** | |
|---|---|---|
| | **aIRR (95%CI)** | **p-value** |
| Age | | |
| 0-9 | 0.69 (0.13-3.65) | 0.663 |
| 10-19 | 0.88 (0.49-1.64) | 0.688 |
| 20-29 | 1.14 (0.96-1.36) | 0.142 |
| 30-39 | 0.86 (0.76-0.97) | 0.012 |
| 40-49 | 0.90 (0.82-0.99) | 0.032 |
| 50-59 | 1.00 | - |
| 60-69 | 0.89 (0.79-1.00) | 0.047 |
| 70-79 | 0.95 (0.79-1.15) | 0.598 |
| 80+ | 1.24 (0.69-2.27) | 0.446 |
| Sex | | |
| Female | 1.00 | - |
| Male | 1.01 (0.93-1.11) | 0.762 |
| Prefer not to say | 0.53 (0.27-1.05) | 0.061 |
| Ethnicity | | |
| White | 1.00 | - |
| Mixed/Multiple ethnic groups | 1.08 (0.76-1.56) | 0.667 |
| Asian/Asian British | 0.53 (0.35-0.81) | 0.003 |
| Black/African/Carribean/Black British | 0.46 (0.17-1.23) | 0.135 |
| Other ethnic group | 0.53 (0.17-1.77) | 0.290 |
| Nation | | |
| England | 1.00 | - |
| Northern Ireland | 0.92 (0.59-1.47) | 0.708 |
| Scotland | 0.79 (0.67-0.94) | 0.006 |
| Wales | 1.23 (1.00-1.51) | 0.052 |
| Household size | | |
| 1 | 1.00 | - |
| 2 | 0.92 (0.83-1.02) | 0.133 |

| | Multivariable analysis[1] | |
|---|---|---|
| | aIRR (95%CI) | p-value |
| 3 | 1.00 (0.89-1.13) | 0.981 |
| 4 | 0.85 (0.75-0.96) | 0.010 |
| 5 | 0.98 (0.83-1.17) | 0.839 |
| 6+ | 0.88 (0.68-1.16) | 0.357 |
| Day of the week (contacts recorded) | | |
| Weekday | 1.00 | - |
| Weekend | 1.14 (0.99-1.31) | 0.074 |
| Left home yesterday | | |
| No | 1.00 | |
| Yes | 5.60 (4.94-6.36) | <0.001 |
| School or work situation | | |
| Employed - working from home | 1.00 | - |
| School pupil - studying at home | 0.84 (0.29-2.48) | 0.752 |
| School pupil - attending school | 2.96 (0.97-10.23) | 0.074 |
| College or University student | 0.86 (0.65-1.14) | 0.287 |
| Employed - going to place of work | 3.32 (3.02-3.66) | <0.001 |
| Self Employed | 1.63 (1.43-1.87) | <0.001 |
| Healthcare professional | 5.05 (4.25-6.03) | <0.001 |
| Unemployed | 1.11 (0.90-1.37) | 0.354 |
| Furloughed | 1.20 (0.98-1.48) | 0.074 |
| Unable to work | 0.96 (0.71-1.30) | 0.798 |
| Retired | 1.24 (1.09-1.42) | 0.001 |
| COVID-19 circumstance | | |
| Not self-isolating or shielding | 1.00 | - |
| Self-isolating - I have symptoms* | 4.07 (1.96-9.79) | 0.001 |
| Self-isolating - Someone in my household has symptoms | 1.30 (0.51-3.60) | 0.614 |
| Self-isolating - Someone in my support bubble has symptoms | 0.82 (0.21-3.34) | 0.780 |
| Self-isolating - precaution/told to do so by Test and Trace | 0.58 (0.43-0.79) | <0.001 |
| Shielding - I am a vulnerable individual | 0.81 (0.66-1.01) | 0.050 |

| | Multivariable analysis[1] | |
|---|---|---|
| | aIRR (95%CI) | p-value |
| Shielding - I live with a vulnerable individual | 0.79 (0.62-1.01) | 0.063 |
| Not sure | 0.55 (0.39-0.78) | 0.001 |

[1]Adjusted for age, sex, ethnicity, nation, household size, day of the week, left home, school or work situation and COVID-19 circumstance.
*This increased contact rate is due to one participant who was self-isolating with symptoms reporting a high number of contacts (*see results*).

**Table S8.** Association of participant characteristics with risk (adjusted odds ratios) of visiting another household (N = 4,030). Model intercept 0.11 (0.09-0.14).

| | **Adjusted Odds Ratio (95%CI)**[1] | **p-value** |
|---|---|---|
| Age | | |
| 10-19 | 0.75 (0.18-2.20) | 0.643 |
| 20-29 | 0.77 (0.47-1.21) | 0.277 |
| 30-39 | 0.85 (0.62-1.15) | 0.313 |
| 40-49 | 1.03 (0.82-1.30 | 0.805 |
| 50-59 | 1.00 | - |
| 60-69 | 1.09 (0.86-1.38) | 0.479 |
| 70-79 | 1.18 (0.79-1.72) | 0.413 |
| 80+ | 0.73 (0.11-2.73) | 0.682 |
| Sex | | |
| Female | 1.23 (0.98-1.56) | 0.073 |
| Male | 1.00 | - |
| Prefer not to say | 1.07 (0.17-3.92) | 0.926 |
| Part of a support bubble | | |
| No | 1.00 | - |
| Yes | 1.92 (1.61-2.28) | <0.001 |

[1]Adjusted for age, sex and whether a participants was a part of a support bubble.
*0-9 year olds excluded from analysis due to insufficient data.

**Survey Questions - Round 1**

Q1
Are you aged 13 or over?

◯ Yes

◯ No

Q1.a
**Please make sure you agree to the following before continuing with the survey:**
You currently live in the UK;
You have read the Participant Information Sheet and fully understand what is expected of you within this study;
Your participation is voluntary and you are aware that you can stop the survey at any point; You understand that data submitted prior to closing the survey will be collected;
You consent to Lancaster University keeping the anonymised data for a period of 10-years after the study has finished;
If you are filling out the survey on behalf of someone else, please make sure you have their consent before continuing.

◯ **I consent to taking part in the CoCoNet study**

Q1.b
**If you are under the age of 13 we do ask that a parent or guardian fills out the survey with you.**

Please take the time to read through and discuss the *Information sheet for Children* together. The parent or guardian should also read through the more detailed *Participant Information Sheet*.

**Please make sure you both agree to the following before continuing with the survey:**
I live in the UK;
I have read and understood the information sheet(s);
I understand I can stop the survey at any point;
I understand that my answers will be kept for 10 years after the study has finished.

| | Child | Parent/Guardian |
|---|---|---|
| **I agree to take part in the CoCoNet study/ I consent to my child taking part in the study** | ☐ | ☐ |

Q2 Where in the UK do you currently live?

- ◯ England
- ◯ Northern Ireland
- ◯ Scotland
- ◯ Wales
- ◯ I do not live in the UK

Q3 What is your age?

- ◯ 0 - 9 years old
- ◯ 10 - 19 years old
- ◯ 20 - 29 years old
- ◯ 30 - 39 years old
- ◯ 40 - 49 years old
- ◯ 50 - 59 years old
- ◯ 60 - 69 years old
- ◯ 70 - 79 years old
- ◯ Aged 80 or over

Q4 What is your sex?
*The answer you give can be different from what is on your birth certificate.*

- ◯ Female
- ◯ Male
- ◯ Prefer not to say

Q5 Which of the following best describes your ethnicity?

- ◯ English / Welsh / Scottish / Northern Irish / British
- ◯ Irish
- ◯ Gypsy or Irish Traveller
- ◯ Any other White background
- ◯ White and Black Caribbean
- ◯ White and Black African
- ◯ White and Asian
- ◯ Any other Mixed / Multiple ethnic background
- ◯ Indian
- ◯ Pakistani
- ◯ Bangladeshi
- ◯ Chinese
- ◯ Any other Asian background
- ◯ African
- ◯ Caribbean
- ◯ Any other Black / African / Caribbean background
- ◯ Arab
- ◯ Any other ethnic group
- ◯ Prefer not to say

Q6 What is the first part of your home postcode?
*For example, if your home postcode was LA1 4YW then you would enter LA1.*

______________________________________________________________

Q7 Which type of accommodation best describes your home?

- ◯ Flat, maisonette or apartment
- ◯ House or bungalow
- ◯ Mobile or temporary structure
- ◯ Assisted living facility
- ◯ Care home
- ◯ Other

Q8
What is your current school or work situation?

- ◯ School pupil - studying at home
- ◯ School pupil - still attending school
- ◯ College or University student
- ◯ Employed - working from home
- ◯ Employed - still going to place of work
- ◯ Self Employed
- ◯ Healthcare professional
- ◯ Unemployed
- ◯ Furloughed
- ◯ Unable to work
- ◯ Retired
- ◯ Other

Q9 Currently, do you regularly meet members of the general public as part of your job?

- ◯ Yes
- ◯ No

Q10
Are you self-isolating or shielding because of COVID-19?
*A vulnerable individual here refers to a clinically extremely vulnerable person.*

- ○ I am not self-isolating or shielding
- ○ Self Isolating - I have symptoms of COVID
- ○ Self Isolating - Someone in my household has symptoms of COVID
- ○ Self Isolating - Someone in my support bubble has symptoms of COVID
- ○ Self Isolating - As a precaution / told to do so by Test and Trace
- ○ Shielding - I am a vulnerable individual
- ○ Shielding - I live with a vulnerable individual
- ○ Not sure

Q11
How many other people currently live with you at home?

- ○ 0 - I live alone
- ○ 1
- ○ 2
- ○ 3
- ○ 4
- ○ 5 or more

Q12

How many people of each age group live with you at home?

*Do not include yourself.*

*Drop down options of 0, 1, 2, 3, 4, 5 or more for each age group.*

0 - 9 year olds
10 - 19 year olds
20 - 29 year olds
30 - 39 year olds
40 - 49 year olds
50 - 59 year olds
60 -69 year olds
70 - 79 year olds
Aged 80 or over

Q13

Have you formed a support bubble with another household?

*A single-person household can join with one other household and interact without maintaining social distance.*

- ○ Yes
- ○ No

Q14

How many people of each age group are part of your support bubble?

*Do not include your own household members.*

*Drop down options of 0, 1, 2, 3, 4, 5 or more for each age group.*

0 - 9 year olds
10 - 19 year olds
20 - 29 year olds
30 - 39 year olds
40 - 49 year olds
50 - 59 year olds
60 -69 year olds
70 - 79 year olds
Aged 80 or over

Q15

Thinking about the past 7 days, on how many of these days did you meet someone from your support bubble?

- ◯ None
- ◯ 1 day
- ◯ 2 days
- ◯ 3 days
- ◯ 4 days
- ◯ 5 days
- ◯ 6 days
- ◯ 7 days
- ◯ Not sure

Q16

Did you leave your home or property yesterday?

*Do not include going into your private garden, but do include visits to shared or communal gardens or spaces.*

- ○ Yes
- ○ No

Q17

Where did you go yesterday? Tick all that apply.

- ☐ Visited the home of someone else
- ☐ My school or workplace
- ☐ Doctor's surgery or healthcare facility
- ☐ Supermarket or convenience store
- ☐ Other shops or retail spaces (e.g. garden centre, clothing shops, drive-through food outlets)
- ☐ Restaurant, café or pub
- ☐ For a walk or exercise
- ☐ Other - *please do not include any identifying information*

____________________________________________

Q18

What modes of transport did you use yesterday? Tick all that apply.

- ☐ I walked or cycled
- ☐ I travelled in a car by myself
- ☐ I travelled in a car with another person(s)
- ☐ I took a bus, tram or train
- ☐ I took an aeroplane or ferry

Q19
Not including those that you live with, how many people did you meet yesterday?
*Only include those you had a face-to-face conversation with.*

- ○ None
- ○ 1
- ○ 2
- ○ 3
- ○ 4
- ○ 5
- ○ 6
- ○ 7
- ○ 8
- ○ 9
- ○ 10
- ○ 11
- ○ 12
- ○ 13
- ○ 14
- ○ 15 or more

Q20
Please tell us about each of the people you met yesterday.
*Information collected for up to 14 contacts.*

| | How old were they? *Please estimate the person's age if you are unsure.* | | | | | | | | Did you meet this person indoors? *For example, in a shop, office or house.* | | Did anyone that you live with also meet this person yesterday? | | |
|---|---|---|---|---|---|---|---|---|---|---|---|---|---|
| | 0 - 4 | 5 - 9 | 10 - 19 | 20 - 39 | 40 - 59 | 60 - 69 | 70 - 79 | 80 + | Yes | No | Yes | No | Not sure |

Q21
Please tell us about each of the people you met yesterday.
*Information collected for up to 14 contacts.*

| | How old were they? *Please estimate the person's age if you are unsure.* | | | | | | | | Did you meet this person indoors? *For example, in a shop, office or house*. | |
|---|---|---|---|---|---|---|---|---|---|---|
| | 0 - 4 | 5 - 9 | 10 - 19 | 20 - 39 | 40 - 59 | 60 - 69 | 70 - 79 | 80 + | Yes | No |

Q22
How many people of each age group did you meet yesterday?
*Drop down choice of integers 1 - 19 or '20 or more' for each age group.*

0-4 year olds

5-9 year olds

10-19 year olds

20-39 year olds

40-59 year olds

60-69 year olds

70-79 year olds

Aged 80 or over

Q23
Did you meet these people indoors or outdoors?
*For example, meeting someone indoors could be in a shop, office or house etc.*

- ○ I met everyone indoors
- ○ I met most people indoors
- ○ I met most people outdoors
- ○ I met everyone outdoors
- ○ Not sure

Q24
Were you able to maintain social distance from everyone you met yesterday?
*Do not include people that you live with or those in your support bubble. Please refer to the government advice for the recommended social distance in your area.*

- ◯ Yes, all of the time
- ◯ More than half of the time
- ◯ Less than half of the time
- ◯ No, none of the time
- ◯ Not sure

Q25 Of the people you live with, how many people stayed at home all day yesterday?

- ◯ None
- ◯ 1
- ◯ 2
- ◯ 3
- ◯ 4
- ◯ 5 or more
- ◯ Not sure

Q26 Thinking about the past 7 days, on how many of these days did you leave your home or property?

- ◯ None
- ◯ 1 day
- ◯ 2 days
- ◯ 3 days
- ◯ 4 days
- ◯ 5 days
- ◯ 6 days
- ◯ 7 days
- ◯ Not sure

Q27 What was the furthest distance from home you travelled over the past 7 days?

- ◯ Less than 2 miles (3 km)
- ◯ 2 - 9 miles (3 - 15 km)
- ◯ 10 - 19 miles (16 - 31 km)
- ◯ 20 - 49 miles (32 - 79 km)
- ◯ 50 miles (80 km) or more
